# Improving primary prevention of hypertension using a social media website

**DOI:** 10.1101/2021.06.28.21259064

**Authors:** A.E. Demkina, M.V. Bezzubtseva, M.N. Lipilina, K.S. Benimetskaya, A.L. Pivenstein, N.D. Gavrilyuk, A.V. Isaeva, F.A. Lobzhanidze, N.V. Podgorodetskaya, V.G. Klyashtornyy, V.Yu. Taskina

## Abstract

**Purpose:** To determine the most effective methods of informing people about the primary prevention of hypertension using a social media website.

**Materials and methods:** It was a randomized, prospective, two-stage study conducted on a social media website (*i*.*e*., the Instagram platform). At Stage 1 *Save Your Heart* online school was announced, and 945 applicants were registered for the training. The online training programme included the following modules: Risk Factors for Cardiovascular Diseases (CVDs), Heart-Healthy Diet, Cholesterol. What a Patient Needs to Know, Physical Activity for the Prevention of CVDs, Overweight and Obesity, Smoking as a Risk Factor for CVDs, Hypertension: Diagnosis, Hypertension: Treatment, Myocardial Infarction: Diagnosis and Treatment. At Stage 2 a total of 125 participants were randomly selected and assigned into one of the four training groups depending on the training mode, *i*.*e*. text posts of up to 4.000 characters, short video clips of up to 5 minutes, text posts followed by video clips or video clips followed by text posts in Group 1 (*n*=31), Group 2 (*n*=31), Group 3 (*n*=33) and Group 4 (*n*=30), respectively. Before and after training respondents in all the four groups completed the Heart Disease Knowledge Questionnaire (HDKQ).

**Results:** The total number of people who listened to and read the materials of the online school was 2.108 people; the overall engagement (comments, shares and saves) was 1.598 people. The average percentage viewed was 22%. The online school audience was mostly female (84%). A total of 80.8% of participants had post-secondary education. The mean duration of hypertension was 6.1 years. Smokers accounted for 3.2% of the respondents. Before training the number of correct responses for 29 HDKQ statements was 18.4 (CI, 17.5;19.4), with no significant differences between the groups. After training the number of correct responses increased to 21.9 (CI, 21;22.7) (*p* (ANOVA) <0.0001). The post-hoc analysis showed that after training the respondents from Group 3 gave the lowest number of correct responses compared to Groups 1 and 2, *i*.*e*. Δ=4.9 (CI, -7.8;2.0) and Δ=3.7 (CI, -6.5;-0.8), respectively. Participants from Group 4 had more correct responses than the respondents in Group 3, *i*.*e*. Δ=5.2 (CI, 2.2;8.1). The regression analysis showed that the post-training number of correct responses in Group 4 increased on average by 3.9 compared to Group 3 (β=3.94 *p*=0.04 (CI, 0.21;7.66)). The study showed a significant association between the duration of hypertension and the number of correct responses both before and after training (β=0.20 *p*=0.007 (CI, 0.06;0.34) and β=0.16 *p*=0.005 (CI, 0.05;0.27), respectively). No association was found between gender, age, education and the number of correct responses both before and after training.

**Conclusions:** In all the 4 groups there was a tendency to increase in the number of correct responses after training, but among the training modes the most effective method of informing people about the primary prevention of hypertension using a social media website corresponded to the following sequence: a video clip of up to 5 minutes followed by a text post of up to 4,000 characters. Participants in Group 3 who received the material in the form of text posts followed by video clips gave the lowest number of correct responses for HDKQ statements after training. The results of this study can be used to design online training programmes for the primary prevention of hypertension.

---

In recent years, due to the widespread use in developed countries, social media have been used to improve the health literacy of patients (1, 2). The leading cardiology societies approve of the use of social media for the prevention of cardiovascular diseases (CVDs) and consider it a promising area (3, 4). Instagram is one of the most popular social networks in the Russian Federation and worldwide. In Russia it has more than 44 million users (5). Notably, in 2019 Russia ranked first among European countries in terms of the use of social media with an average daily usage time of more than two hours.

The increasing popularity of social media and mobile health apps has led to the emergence of ‘mobile health’, a term which combines the use of mobile devices and wireless technologies to support health initiatives both as a method of patient follow-up and data collection and as an effective means of health education and promotion (6). According to the eHealth literacy study published in the Journal of Medical Internet Research, not only middle-aged people, but almost 90% of older people use social media to find and share health information (7). The percentage of people in a similar age group using social media in Russia is likely to be less, but the tendency to an increase in the number of social media users in all age groups is clear.

To date, a number of studies are available on the effective use of the Instagram social media platform to conduct training projects for CVD patients in the Russian Federation (8). Unlike many social networks, Instagram can provide patients with information in both text and video form. Naturally, the question arises as to which form of information presentation is more effectively absorbed by patients. Given the importance of hypertension as one of the leading risk factors for CVDs and the high prevalence of this condition, it is feasible to study the effectiveness of methods of informing people with hypertension about the primary prevention of CVDs using the Instagram social media platform.

## Purpose

To determine the most effective ways to inform people with hypertension about the primary prevention of CVDs using a social media website.

## Material and methods

The study protocol was approved by the Ethics Committee of the Research and Practical Clinical Centre for Diagnostics and Telemedicine Technologies of the Moscow Healthcare Department. All participants gave their written informed consent to participate in the study.

The study was conducted on the Instagram social media platform in 2 stages. At Stage 1 *Save Your Heart* free online school was announced on 8 medical blogs: @doc_4_you, @kardiolog_mv, @doc_cardio_podgorodetskaya, @dr.gavriliuk, @doctor_lobzhanidze, @doc.for.health, @dr_cardioann, @doctor_isaeva_cardio.

At Stage 2 from 945 respondents wishing to participate in the study 125 participants were randomly selected and assigned into one of the four training groups using a random number generator:

1. @sohrani.svoe.serdce.t (*n*=31): health education materials of the online school were represented only by text posts of up to 4,000 characters (9 posts in total);
2. @sohrani.svoe.serdce.v (*n*=31): health education materials of the online school were represented by short video clips of up to 5 minutes (9 posts in total);
3. @sohrani.svoe.serdce.tv (*n*=33): health education materials of the online school were represented by text posts followed by video clips (18 posts in total);
4. @sohrani.svoe.serdce.vt (*n*=30): health education materials of the online school were represented by video clips followed by text posts (18 posts in total).

The training started on 1 February 2021. The education materials were prepared by a group of cardiologists running their medical blogs (@doc_4_you, @kardiolog_mv, @doc_cardio_podgorodetskaya, @dr.gavriliuk, @doctor_lobzhanidze, @doc.for.health, @dr_cardioann, @doctor_isaeva_cardio) based on the guidelines of the European Society of Cardiology and approved by an independent ethics committee.

The programme included the following modules:

1. Risk Factors for CVDs.
2. Heart-Healthy Diet.
3. Cholesterol. What a Patient Needs to Know.
4. Physical Activity for the Prevention of CVDs.
5. Overweight and Obesity.
6. Smoking as a Risk Factor for CVDs.
7. Hypertension: Diagnosis.
8. Hypertension: Treatment.
9. Myocardial Infarction: Diagnosis and Treatment.

Posts were made in all the groups simultaneously 1 time a day in the morning. Feedback from cardiologists was available to participants in the comments under the posts. The education materials contained answers to all the questions of the Heart Disease Knowledge Questionnaire (HDKQ) to be completed.

One week before the training start, a png file with an adapted HDKQ with additional questions on gender, age, education, duration of hypertension, smoking status and body mass index (BMI) was sent to all participants in direct messages. The respondents sent their responses in direct messages of the school accounts in the ‘question number-answer’ form. The response options included ‘Yes’, ‘No’, ‘I don’t know’. On the training end date the adapted HDKQ was once again sent to all participants in direct messages; responses were accepted within 2 weeks after the mailing date.

### Statistical data processing

The data analysis included all study subjects for whom the responses to the questionnaire were obtained at least in one time point (before and/or after training).

For each study subject the number of correct responses to the questionnaire was calculated (ranging from the minimum score of 0 to the maximum score of 29 points). Study results were reported using descriptive statistics indicating the number of non-missing values (N), minimum (Min), maximum (Max), mean (M), standard deviation (SD), 95% confidence interval (CI) for the mean, median (Me) and interquartile range (IQR).

The inter-group comparison was made using analysis of variance (ANOVA). Significant post-hoc differences were identified by pairwise comparison using t test adjusted for the multiple comparisons using the Tukey method. The significance level was set at 0.05 (two-tailed). Additionally, regression models were built. The number of correct responses before training, after training, and the difference between the number of correct responses before and after training for each study subject were used as the dependent variable. Gender, age, education (higher or secondary), group, duration of hypertension, history of smoking, height, weight and body mass index (BMI) were used as factors. The regression coefficient (β) and 95% CI for β were estimated for each factor. Statistical analyses were performed using Stata14.

## Results

The total number of people who listened to and read the materials of the *Save Your Heart* online school was 2,108 people; the overall engagement (comments, shares and saves) was 1,598 people. The average percentage viewed was 22%. The online school audience was mostly female (84%), with a mean age of 45.9±12.3 years. A total of 80.8% of participants had post-secondary education. The mean duration of hypertension was 6.1±7.6 years. Smokers accounted for 3.2% of the respondents. Participant characteristics by group are shown in Table 1.

**Table 1.** General characteristics of the study participants by group.

| Parameter |  | 1 (n=31) | 2 (n=31) | 3 (n=33) | 4 (n=30) |
| --- | --- | --- | --- | --- | --- |
| Gender, n/%: | Male | 5/16 | 5/16 | 6/18 | 4/13 |
|  | Female | 26/84 | 26/84 | 27/82 | 26/87 |
| Age, years | | 43.3±13.6 | 45.3±11.4 | 48.5±11.5 | 46.1±12.9 |
| Post-secondary education, n/% |  | 26/84 | 22/71 | 29/89 | 24/80 |
| Duration of hypertension, years | | 3.3±5.1 | 6.5±6.7 | 7.1±8.7 | 7.9±9.4 |
| Smoking, n |  | 1 | 0 | 2 | 1 |
| BMI, kg/m<sup>2</sup> | | 26.0±5.0 | 26.8±5.3 | 28.6±5.8 | 26.5±4.9 |
| Average percentage viewed, % |  | – | 26 | 23.3 | 23.9 |

Before training the number of correct responses for 29 HDKQ statements was 18.4±5.1 (CI, 17.5;19.4), with no significant differences between the groups (Table 2).

**Table 2.** Descriptive statistics for the number of correct responses before training.

| Parameter | Group 1 (n=31) | Group 2 (n=31) | Group 3 (n=33) | Group 4 (n=30) | Total (n=125) |
| --- | --- | --- | --- | --- | --- |
| Mean | 19.5 | 17.6 | 17.4 | 19.3 | 18.4 |
| 95% CI | (17.7;21.4) | (15.8;19.4) | (15.3;19.5) | (17.7;21.0) | (17.5;19.4) |
| Min/Max | (7;29) | (9;26) | (6;26) | (9;27) | (6;29) |
| p50 | 20 | 17 | 17 | 19 | 19 |
| IQR | 8 | 7 | 10 | 7 | 8 |
| *p* (ANOVA) | 0.213 |  |  |  |  |

**Table 3.**
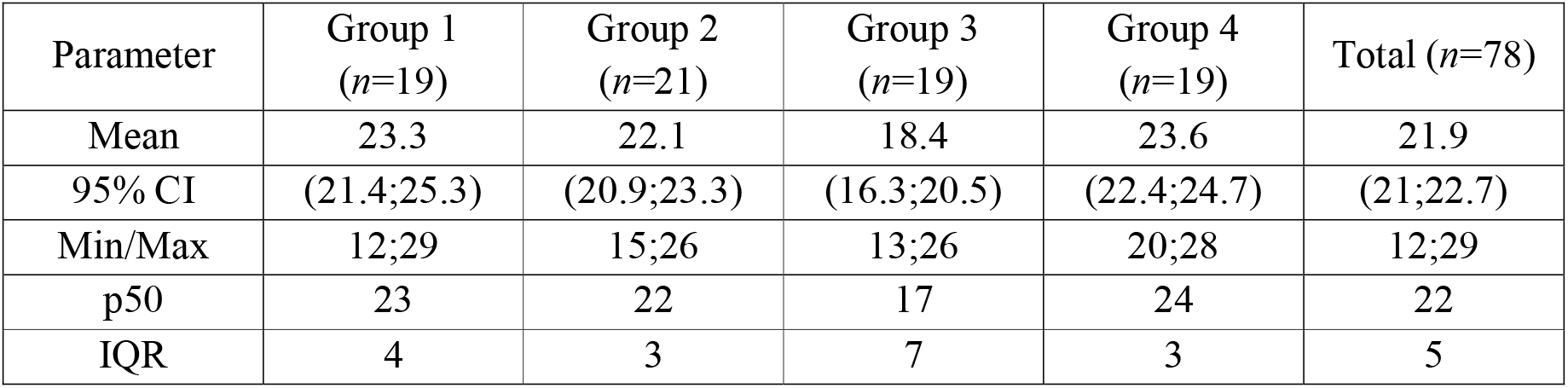
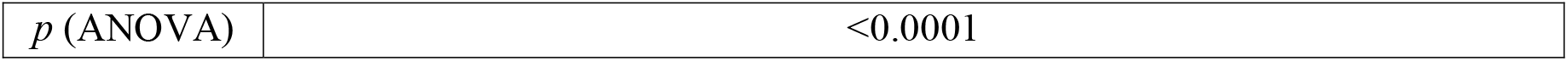
Descriptive statistics for the number of correct responses after training.

| Parameter | Group 1 (n=19) | Group 2 (n=21) | Group 3 (n=19) | Group 4 (n=19) | Total (n=78) |
| --- | --- | --- | --- | --- | --- |
| Mean | 23.3 | 22.1 | 18.4 | 23.6 | 21.9 |
| 95% CI | (21.4;25.3) | (20.9;23.3) | (16.3;20.5) | (22.4;24.7) | (21;22.7) |
| Min/Max | 12;29 | 15;26 | 13;26 | 20;28 | 12;29 |
| p50 | 23 | 22 | 17 | 24 | 22 |
| IQR | 4 | 3 | 7 | 3 | 5 |

|  |  |
| --- | --- |
| $p$ (ANOVA) | <0.0001 |

After training the number of correct responses increased to 21.9±3.9 (CI, 21;22.7) (*p* (ANOVA) <0.0001).

The post-hoc analysis showed that after training the respondents from Group 3 gave the lowest number of correct responses compared to Groups 1 and 2, *i*.*e*. Δ=4.9 (CI, –7.8;2.0) and Δ=3.7 (CI, –6.5;–0.8), respectively. Participants from Group 4 had more correct responses than the respondents in Group 3, *i*.*e*. Δ=5.2 (CI, 2.2;8.1). The regression analysis showed that the post-training number of correct responses in Group 4 increased on average by 3.9 compared to Group 3 (β=3.94 *p*=0.04 (CI, 0.21;7.66) (Table 4).

**Table 4.** Post-hoc pairwise comparison results for the number of correct responses after training.

| Indicator | Mean difference | 95% CI for difference | Significance |
| --- | --- | --- | --- |
| Group 2 vs Group 1 | -1.2 | -4.1;1.6 | – |
| Group 3 vs Group 1 | -4.9 | -7.8;-2.0 | + |
| Group 4 vs Group 1 | 0.3 | -2.6;3.2 | – |
| Group 3 vs Group 2 | -3.7 | -6.5;-0.8 | + |
| Group 4 vs Group 2 | 1.5 | -1.3;4.3 | – |
| Group 4 vs Group 3 | 5.2 | 2.2;8.1 | + |

The study showed a significant association between the duration of hypertension and the number of correct responses both before and after training (β=0.20 *p*=0.007 (CI, 0.06;0.34) and β=0.16 *p*=0.005 (CI, 0.05;0.27), respectively). No association was found between gender, age, education and the number of correct responses both before and after training (Tables 5, 6).

**Table 5.** Regression parameters for the number of correct responses before training (*N*=98)

| Factor | $\beta$ | $p$ | 95% CI for $\beta$ |
| --- | --- | --- | --- |
| Gender (Female = ref) | -0.50 | 0.760 | -3.73;2.74 |
| Age | -0.05 | 0.270 | -0.13;0.04 |
| Education (Secondary = ref) | -0.05 | 0.970 | -2.72;2.62 |
| Group (3 = ref) |  |  |  |
| 1 | 1.76 | 0.198 | -0.94;4.46 |
| 2 | 0.37 | 0.786 | -2.33;3.07 |
| 4 | 0.83 | 0.551 | -1.92;3.58 |
| Duration of hypertension | 0.20 | 0.007 | 0.06;0.34 |
| History of smoking | 0.43 | 0.011 | 0.10;0.76 |
| Height | 1.44 | 0.972 | -80.9;83.79 |
| Weight | -0.04 | 0.932 | -0.93;0.85 |
| BMI | -0.15 | 0.905 | -2.63;2.33 |
| Intercept | 23.84 | 0.731 | -113.78;161.47 |

**Table 6.** Regression parameters for the number of correct responses after training (*N*=70)

| Factor | $\beta$ | $p$ | 95% CI for $\beta$ |
| --- | --- | --- | --- |
| Gender (Female = ref) | -0.33 | 0.823 | -3.28;2.62 |
| Age | -0.07 | 0.051 | -0.14;0.00 |
| Education (Secondary = ref) | 1.70 | 0.176 | -0.78;4.18 |
| Group (3 = ref) |  |  |  |
| 1 | 3.67 | 0.005 | 1.18;6.15 |
| 2 | 2.33 | 0.062 | -0.12;4.78 |
| 4 | 3.34 | 0.011 | 0.79;5.90 |
| Duration of hypertension | 0.16 | 0.005 | 0.05;0.27 |
| History of smoking | 0.08 | 0.624 | -0.23;0.39 |
| Height | -9.23 | 0.798 | -80.92;62.46 |
| Weight | 0.12 | 0.768 | -0.67;0.90 |
| BMI | -0.42 | 0.702 | -2.63;1.78 |
| Intercept | 38.78 | 0.520 | -81.24;158.81 |

## Discussion

In this study the most effective way to inform people with hypertension about the primary prevention of CVDs on social media was the provision of information in the form of a video clip of up to 5 minutes followed by a text post of up to 4,000 characters.

Currently, the WHO recognises social resources as a valuable agent for behaviour change in health promotion (9). The development of digital technologies and the ever-increasing usage of social media determine their potential to provide information and social support for people with various non-communicable diseases (10–12). This is why social media are increasingly being used as an educational resource in healthcare (13). The use of social media for learning and education has already proven effective in obesity and diabetes mellitus (14,15).

In a recent study published in Surgery 95% of respondents reported that joining the Facebook support group for liver transplant patients had a positive impact on their care (16).

However, despite the large amount of health information on social media (13,17,18), we do not know much about the comprehensibility of the presented material. Thus, it is important to study various ways of education material presentation.

As shown in this study, the post-training number of correct responses increased in all study groups. Among the training modes we studied, the most effective method of informing people about the primary prevention of hypertension using a social media website corresponded to the following sequence: a video clip of up to 5 minutes followed by a text post of up to 4,000 characters. The lowest number of correct responses for HDKQ statements after training was given by the participants who received the material in the form of text posts followed by video clips.

It is known that text- and video-based patient education is effective in the short-term treatment of out- and inpatients (19). At the same time, the effectiveness of education through social media was studied in a limited number of patients with obesity and diabetes mellitus (20). In this study, different modes of hypertensive patient education on social media were compared for the first time.

The number of correct responses before and after training depended on the duration of hypertension, which is consistent with a number of studies (21); however, not all studies established this association (22). In this study gender, age, smoking status and body mass index were not associated with the awareness of risk factors.

Thus, the results obtained in the *Save Your Heart* school for hypertensive patients confirm the possibility of effective training of patients in the primary prevention of CVDs in the short term provided that a combined programme is used in a certain sequence (video clips followed by texts).

## Conclusions

Thus, as shown in this study, in all the 4 groups there was a tendency to increase in the number of correct responses after training, but among the training modes the most effective method of informing people about the primary prevention of hypertension using a social media website corresponded to the following sequence: a video clip of up to 5 minutes followed by a text post of up to 4,000 characters. Participants in Group 3 who received the material in the form of text posts followed by video clips gave the lowest number of correct responses for HDKQ statements after training. The results of this study can be used to design online primary prevention training programmes for people with hypertension.

## Data Availability

Absent

## References

1. Dermatology on Instagram: An Analysis of Hashtags - PubMed. Accessed April 29, 2021. https://pubmed.ncbi.nlm.nih.gov/29601627/

2. Eshah NF. Investigating cardiovascular patients’ preferences and expectations regarding the use of social media in health education. Contemp Nurse. 2018;54(1):52–63. doi:10.1080/10376178.2018.1444497

3. Rumsfeld JS, Brooks SC, Aufderheide TP, et al. Use of mobile devices, social media, and crowdsourcing as digital strategies to improve emergency cardiovascular care. Circulation. 2016;134(8):e87–e108. doi:10.1161/CIR.0000000000000428

4. Parwani P, Choi AD, Lopez-Mattei J, et al. Understanding Social Media: Opportunities for Cardiovascular Medicine. J Am Coll Cardiol. 2019;73(9):1089–1093. doi:10.1016/j.jacc.2018.12.044

5. Social media in Russia - Statistics & Facts | Statista. Accessed April 9, 2021. https://www.statista.com/topics/6281/social-media-in-russia/

6. Nayak BS, Lewis LE, Margaret B, Bhat Y R, D’Almeida J, Phagdol T. Randomized controlled trial on effectiveness of mHealth (mobile/smartphone) based Preterm Home Care Program on developmental outcomes of preterms: Study protocol. J Adv Nurs. 2019;75(2):452–460. doi:10.1111/jan.13879

7. Tennant B, Stellefson M, Dodd V, et al. eHealth literacy and Web 2.0 health information seeking behaviors among baby boomers and older adults. J Med Internet Res. 2015;17(3). doi:10.2196/jmir.3992

8. Demkina AE, Ryabinina MN, Aksenova GA, et al. Testing the educational program “primary and secondary prevention of cardiovascular diseases” on the basis of social networking service Instagram. Russ J Cardiol. 2020;25(9):13–19. doi:10.15829/1560-4071-2020-3932

9. World Health Organisation. Regional Office for Europe. Health 2020: a European policy framework supporting action across government and society for health and well-being. Accessed April 29, 2021. http://www.euro.who.int/PubRequest?language=Russian.

10. Patel R, Chang T, Greysen SR, Chopra V. Social media use in chronic disease: A systematic review and novel taxonomy. Am J Med. 2015;128(12):1335–1350. doi:10.1016/j.amjmed.2015.06.015

11. Waring ME, Jake-Schoffman DE, Holovatska MM, Mejia C, Williams JC, Pagoto SL. Social Media and Obesity in Adults: a Review of Recent Research and Future Directions. Curr Diab Rep. 2018;18(6):1–9. doi:10.1007/s11892-018-1001-9

12. Gabarron E, Bradway M, Fernandez-Luque L, et al. Social media for health promotion in diabetes: Study protocol for a participatory public health intervention design. BMC Health Serv Res. 2018;18(1). doi:10.1186/s12913-018-3178-7

13. Pizzuti AG, Patel KH, McCreary EK, et al. Healthcare practitioners’ views of social media as an educational resource. Houwink EJF, ed. PLoS One. 2020;15(2):e0228372. doi:10.1371/journal.pone.0228372

14. Abedin T, Al Mamun M, Lasker MAA, et al. Social Media as a Platform for Information About Diabetes Foot Care: A Study of Facebook Groups. Can J Diabetes. 2017;41(1):97–101. doi:10.1016/j.jcjd.2016.08.217

15. Jane M, Hagger M, Foster J, Ho S, Pal S. Social media for health promotion and weight management: A critical debate. BMC Public Health. 2018;18(1). doi:10.1186/s12889-018-5837-3

16. Dhar VK, Kim Y, Graff JT, et al. Benefit of social media on patient engagement and satisfaction: Results of a 9-month, qualitative pilot study using Facebook. Surg (United States). 2018;163(3):565–570. doi:10.1016/j.surg.2017.09.056

17. Al Mamun M, Ibrahim HM, Turin TC. Social media in communicating health information: An analysis of Facebook groups related to hypertension. Prev Chronic Dis. 2015;12(1). doi:10.5888/pcd12.140265

18. Ladeiras-Lopes R, Baciu L, Grapsa J, et al. Social media in cardiovascular medicine: a contemporary review. Eur Hear J - Digit Heal. 2020;1(1):10–19. doi:10.1093/ehjdh/ztaa004

19. Adam M, McMahon SA, Prober C, Bärnighausen T. Human-centered design of video-based health education: An iterative, collaborative, community-based approach. J Med Internet Res. 2019;21(1). doi:10.2196/12128

20. Alanzi T. Role of social media in diabetes management in the middle east region: Systematic review. J Med Internet Res. 2018;20(2). doi:10.2196/jmir.9190

21. Lugo-Mata ¡R, Urich-Landeta AS, Andrades-Pérez AL, León-Dugarte Mj, Marcano-Acevedo LA, Jofreed López Guillen MH. Factors associated with the level of knowledge about hypertension in primary care patients. Med Univ. 2017;19(77):184–188. doi:10.1016/j.rmu.2017.10.008

22. Asiri AA, Asiri S, Asiri H. Knowledge Related to Hypertension Risk Factors, Diet, and Lifestyle Modification: A Comparative Study Between Hypertensive and Non-Hypertensive Individuals. Cureus. 2020;12(8). doi:10.7759/cureus.9890

